# The German Multicenter Registry for ME/CFS (MECFS-R)

**DOI:** 10.1101/2024.04.25.24306335

**Authors:** Hannah Hieber, Rafael Pricoco, Katrin Gerrer, Cornelia Heindrich, Katharina Wiehler, Lorenz L. Mihatsch, Matthias Hägele, Daniela Schindler, Quirin Donath, Catharina Christa, Annika Grabe, Alissa Kircher, Ariane Leone, Yvonne Müller, Hannah Zietemann, Helma Freitag, Franziska Sotzny, Cordula Warlitz, Silvia Stojanov, Daniel B. R. Hattesohl, Anna Hausruckinger, Kirstin Mittelstrass, Carmen Scheibenbogen, Uta Behrends

**Affiliations:** Technical University of Munich, Germany; TUM School of Medicine and Health, Children’s Hospital, Pediatrics, MRI Chronic Fatigue Center for Young People (MCFC); Institut für Med. Immunologie, Immundefektambulanz, Charité – Universitätsmedizin Berlin; Campus Virchow-Klinikum (CVK), Berlin, Germany; Technical University of Munich, Germany; TUM School of Medicine and Health, Children’s Hospital, Child and Adolescent Psychosomatics, MRI Chronic Fatigue Center for Young People (MCFC); German Association for ME/CFS, 20146 Hamburg, Germany; German Center for Infection Research (DZIF), Munich, Germany

**Keywords:** Myalgic encephalomyelitis, chronic fatigue syndrome, ME/CFS, post-viral syndrome, registry, post-COVID, PASC, children, adolescents

## Abstract

Myalgic encephalomyelitis/chronic fatigue syndrome (ME/CFS) is a debilitating multi-systemic disease characterized by a complex, incompletely understood etiology. To facilitate future clinical and translational research, a multicenter German ME/CFS registry was established to collect comprehensive, longitudinal, clinical, epidemiological, and laboratory data from adults, adolescents, and children in a web-based multilayer-secured database.

Here, we present the research protocol and first results of a pilot cohort of 174 ME/CFS patients diagnosed at two specialized tertiary fatigue centers, including 130 (74.7%) adults (mean age 38.4; SD 12.6) and 43 (25.3%) pediatric patients (mean age 15.5; SD 4.2). A viral trigger was identified in 160/174 (92.0%) cases, with SARS-CoV-2 in almost half of them. Patients exhibited severe functional and social impairment, as reflected by a median Bell Score of 30.0 (IQR 30.0 to 40.0) and a poor health-related quality of life assessed with the Short form-36 health survey, resulting in a mean score of 40.4 (SD 20.6) for physical function and 59.1 (SD 18.8) for mental health.

The MECFS-R provides important clinical information on ME/CFS to research and healthcare institutions and, together with a multicenter ME/CFS biobank, will pave the way for research projects addressing the pathogenesis, diagnostic markers, and treatment options.

Trial registration: ClinicalTrials.gov NCT05778006.

## INTRODUCTION

Myalgic encephalomyelitis/chronic fatigue syndrome (ME/CFS) is a frequent, complex, severe, chronic disease classified by the World Health Organization as a neurological disorder (ICD-10 GM G93.3, ICD-10 CM G93.32, ICD-11 8E49) [1].

The reported global prevalence of ME/CFS ranged from 0.2% (clinically diagnosed) to 3.5% (self-reported), depending on the study design and diagnostic criteria applied [2]. In Germany, the pre-pandemic number of affected people is estimated as 140,000 – 310,000, including up to 90,000 children and adolescents at the age of 6-17 years [3, 4]. By definition, patients with ME/CFS suffer from long-lasting symptoms, and only 5% of adult patients experience remission of the disease [5]. A Norwegian population-based study found two age peaks at age 10–19 and 30–39 years [6].

The clinical picture is characterized by a substantial loss in pre-illness levels of activity with pathological exhaustion (fatigue) and long-term worsening of symptoms after mild to moderate activities (post-exertional malaise, PEM) (“crashes”). Fatigue and PEM are typically accompanied by sleep disturbances, pain, cognitive, autonomic, neuroendocrine, and flu-like symptoms [7]. Participation in social life is often severely impaired, and significant absences from school or work are frequent [2, 5].

A febrile episode with confirmed or probable viral origin is usually found before symptom onset. Epstein-Barr virus (EBV)-associated infectious mononucleosis (IM) is a prominent trigger [8] and accounted for about half of the pre-pandemic post-infectious ME/CFS cases in childhood and adolescence [9-13]. In a study in Chicago, 13%, 7%, and 4% adolescents were diagnosed with ME/CFS at 6, 12, and 24 months after EBV-IM [14]. During the pandemic coronavirus disease 2019 (COVID-19) due to infection with the severe acute respiratory syndrome coronavirus 2 (SARS-CoV-2) became the most frequent trigger. Prior research indicates that 19–58% adult outpatients with post-acute sequelae of COVID-19 (PASC) may meet ME/CFS criteria [15-18], and first cases of ME/CFS in children and adolescents with PASC were described [19], though comprehensive population-based studies are currently lacking. The number of ME/CFS cases was expected to at least double during the pandemic due to long-term COVID-19 sequelae [20].

The pathophysiology of ME/CFS is still largely unknown, and reliable biomarkers and specific treatment options are not available yet [21]. Various immunological changes [22-24], including autoantibodies [25, 26], as well as metabolic, vascular dysfunction, and various genetic signatures, have been described [27-29]. Furthermore, persistent or reactivated viruses might contribute to pathogenic mechanisms [30, 31].

ME/CFS is diagnosed by clinical criteria that require PEM as a cardinal symptom. Recommended case definitions include the Canadian Consensus Criteria (CCC) [32] and the broader Institute of Medicine (IOM) criteria [33]. The clinical diagnostic worksheet by Rowe and colleagues (CDW-R) or the pediatric case definition by Jason and colleagues (PCD-J) are being used as age-adapted alternatives for children and adolescents [2, 19, 34]. All case definitions are based on significant severity and frequency of typical ME/CFS symptoms and no evidence of other medical courses, necessitating a thorough diagnostic work-up.

Routine treatment of ME/CFS is symptom-oriented [7]. [7]. It aims at reducing the symptom load with pain, orthostatic intolerance and sleep-related problems, and also the impact of aggravating conditions such as infections, allergies, and/or nutritional deficiencies [35-37]. A key part of managing ME/CFS is the implementation of adequate stress and energy self-management (pacing) to avoid PEM and a subsequent worsening of symptoms. Psychosocial support can help with the development of coping strategies [32]. Providing a timely diagnosis can reduce the complex burden on patients and their social networks and thereby support recovery. Many patients are under- or misdiagnosed and exposed to stigmatization and/or mistreatment [38, 39].

To facilitate future ME/CFS research and to pave the way to improved clinical care, we aimed at a standardized multicenter evaluation of ME/CFS-specific clinical phenotypes and health care features in our novel German ME/CFS registry (MECFS-R). Here, we present the structure of this registry and provide medical data on a cohort of adults, adolescents, and children recruited from two specialized tertiary care centers in Berlin and Munich. The MECFS-R provides comprehensive information on clinical phenotypes, features of medical care, and disease trajectories over time. Together with our ME/CFS biobank, the MECFS-R is expected to aid scientists in discovering risk factors, predictive and diagnostic biomarkers, as well as therapeutic targets for this debilitating disease. It aims to classify distinct patient groups and provide decision-makers with information on the disease’s burden and its social and economic impacts. We will invite additional healthcare providers caring for ME/CFS patients to share our standard diagnostic procedures and contribute data to this registry study.

## METHODS

### Participating Institutions and Target Population

The multicentric MECFS-R was developed by a multidisciplinary team of clinicians, researchers, patients, and members of ME/CFS foundations and support groups. It was established at the Munich Chronic Fatigue Center for Young People (MCFC) in Munich [40] and the Charité Fatigue Center (CFC) in Berlin, Germany [40, 41]. At the CFC most patients are seen in the Department of Immunology with currently approximately 500-600 adult patients seen per year with suspected infection-triggered ME/CFS. In approximately 60% of cases the diagnosis ME/CFS is confirmed. The MCFC sees about 100 young people aged up to 20 years. Additional centers are currently being integrated to create a comprehensive national registry. Standard operation procedures (SOPs) for differential diagnostic workups have been consented, implemented at both centers, and are being distributed nation-wide. Inclusion criteria of the MECFS-R require the diagnosis of ME/CFS by PEM-based clinical criteria (CCC, IOM, CDW-R, PCD-J) and written informed consent provided by the patients or their legal guardians. Exclusion criteria are no ME/CFS diagnosis or missing informed written consent. Participating institutions are collecting detailed clinical routine data for each patient at a baseline and any follow-up visit and storing biosamples, including serum and peripheral blood mononuclear cells (PBMC). The MECFS-R study is registered via ClinicalTrials.gov (NCT05778006).

### Ethical Considerations

Before inclusion into the registry, all patients and/or, in the case of children and adolescents younger than 18 years, their legal guardians, provided written informed consent. The study was approved by the Ethics Committee of the Technical University of Munich Medical Center (MRI TUM), Germany (116/21 S) and by the Ethics Committee of Charité – Universitaetsmedizin Berlin (EA/006/22).

### Data Entry System

The MECFS-R database was built upon the open-source data integration system (DIS) which was initially developed within the Leading Edge Cluster m^4^ funded by the German Federal Ministry of Education and Research (BMBF) [42], is being provided by the Bitcare company, Munich, and is currently hosting various cohorts, including the transplant cohort of the German center of Infection research (DZIF) [43]. DIS offers a secure identity management component and functionality to manage observational data and biosamples. It allows for the integration of data from different digital sources and for advanced security measures such as two-tier pseudonymization, data-at-rest and data-in-transit encryption, role-based access, and audit trails. The ethics and data protection concepts of the DIS have been approved by the relevant local review boards and are in line with the policies of the Data Integration for Future Medicine (DIFUTURE) consortium safeguarding data use and sharing [44]. Following these data protection concept guidelines, a central web-based DIS instance was established at the MRI TUM. Several clinical report forms (CRF) were implemented, encompassing a comprehensive set of routine clinical data from baseline and follow-up visits and providing technical information about stored biosamples. A MECFS-R user manual was developed to facilitate data entry and provide use and access rules. A warning system was introduced into the DIS to inform the user about potential errors. A research coordinator at the MCFC is monitoring completeness of study data and offers online training for each new member of participating sites.

### Data Protection

The data protection concept of the MECFS-R is based on the relevant concepts of the Technology, Methods, and Infrastructure (TMF) for Networked Medical Research. Standard or state-of-the-art IT security measures are used to protect the IT systems. The storage and management of identifying and medical data occurs in separate database systems, which remain organizationally and geographically separate, and pseudonymized in two stages. This separation ensures that any person would need to gain unauthorized access to at least three spatially and organizationally separate subsystems to obtain medical and identifying data. A role-based, personal access authorization system is used to access the registry and managed by the local IT manager. Any data transfer is strictly contingent upon obtaining explicit consent declarations by the registry team and may occur in anonymized or pseudonymized form to safeguard privacy and confidentiality. All data receiving researchers must comply with strict data protection measures and sign an appropriate data usage agreement. Any access to data is strictly project-related. The data stored in the registry may also be used for future research projects approved by the relevant ethics committee. Patients must explicitly consent to the use of their data for other studies. Data is stored for 20 years after completion of the registry. The MRI TUM, represented by its board members, and the participating centers are responsible for data processing. Patients already enrolled will be informed about the multicenter rolling-out process and the inclusion of additional centers in the registry. When turning 18 years of age, participants who were enrolled with the consent of their legal guardians as children or adolescents are informed about their study participation and invited to contact the study center.

### Clinical Phenotyping

Since reliable diagnostic markers are still missing, diagnosing ME/CFS relies on a careful evaluation of the patient’s medical history and clinical symptoms and an appropriate differential diagnostic work-up to exclude alternative causes of the complaints. Comprehensive medical information was derived from our patients by semi-structured interviews on the medical history, including comorbidities, prior diagnostic work-up and prior treatment, as well as by detailed physical examination, psychosocial evaluation, functional and imaging tests, and standardized routine blood analyses. Multiple questionnaires were used to assess individual symptoms, disease severity, health-related quality of life (HRQoL) and participation, including patient-reported outcome measures (PROMs). The presence, severity, and duration of PEM was evaluated by the well-established DePaul Symptom Questionnaire for PEM (DSQ-PEM) [34]. The frequency and severity of ME/CFS symptoms were assessed in a quantitative manner using the 5-point Likert scale provided by our novel Munich Berlin Symptom Questionnaire (MBSQ) [45]. Using the MBSQs diagnostic algorithms, up to four sets of internationally established diagnostic criteria were evaluated, including the CCC and IOM criteria, recommended by the European Network on ME/CFS (EUROMENE) [46] and the Centers for Disease Control and Prevention (CDC) [47] as well as, in the case of children and adolescents, the age-adapted CDW-R [48] and the PCD-J [49]. All of these diagnostic scores required PEM, as internationally recommended. **Table 1** provides an overview of the data sets registered. To answer basic research questions, a minimal core dataset (level 1) was defined for participating primary and secondary care institutions with very limited resources. This level 1 only requires data on age, sex, body mass index (BMI), clinical scores used to establish the ME/CFS diagnosis, duration of PEM, type of trigger, and Bell-Score. To provide data for more comprehensive research questions, a more detailed dataset (level 2) is being offered to tertiary care centers. Contributing centers can apply for site-specific extension of the minimal level 1 or 2 dataset to reflect site-specific standards for routine care and to allow for site-specific evaluations. However, to best avoid missing data in cross-center analyses, any partner site has to agree in providing a complete data set at level 1 or 2.

**Table 1.**
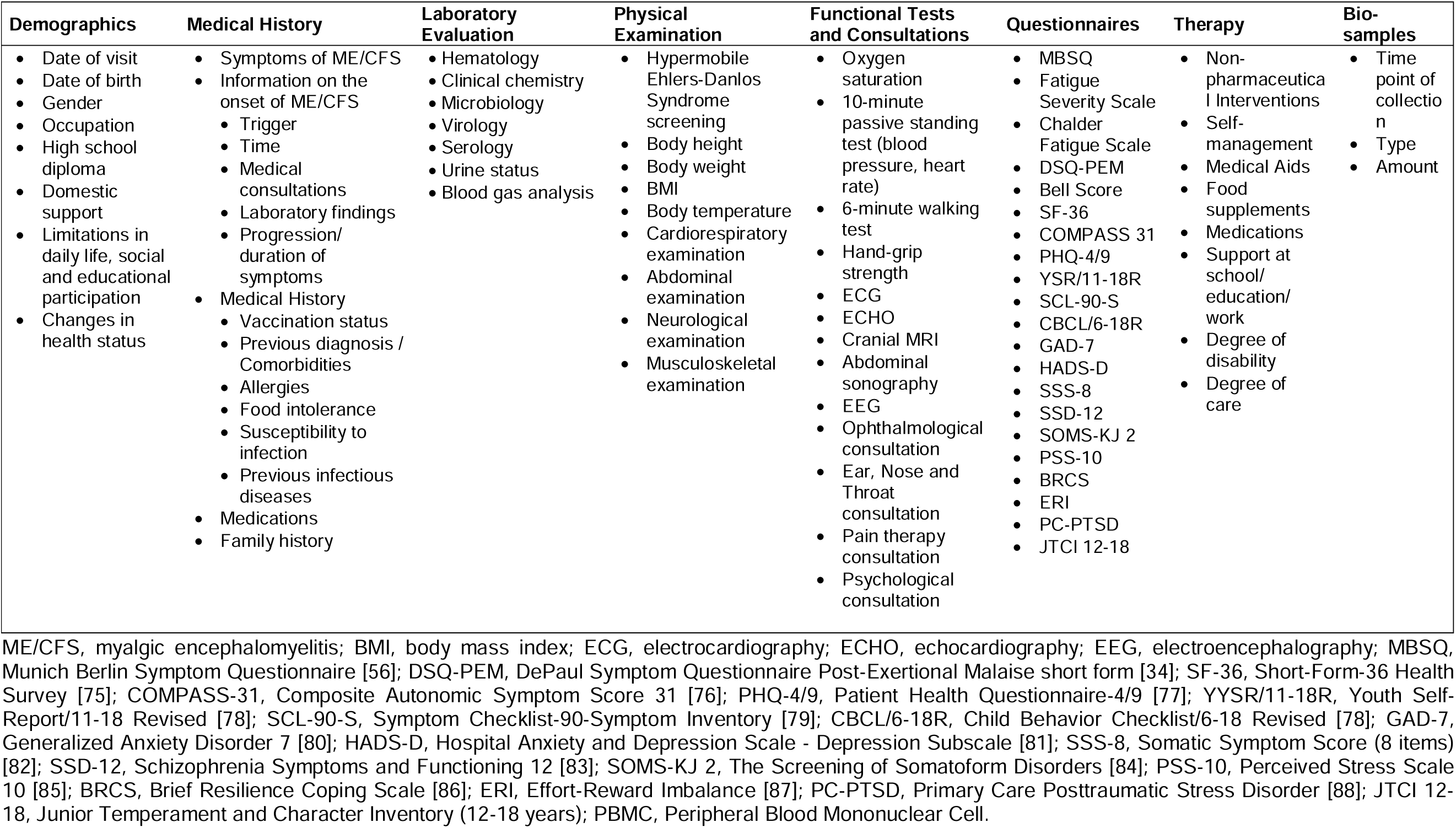
Overview of the Data Collected for the German ME/CFS Registry.

### Patient-Reported Outcomes Measures

The short form-36 health survey (SF-36) is a cross-disease measurement tool to assess HRQoL with good internal consistency and discriminatory validity [50], consisting of 36 items to assess eight dimensions of subjective health: physical functioning, physical role functioning, bodily pain, general health perception, vitality, social functioning, emotional role functioning, and mental well-being, which can be categorized into the fundamental dimensions of physical and mental health. Scores range from 0 (most severe health impairment possible) to 100 points (no health restriction at all). The Bell Score is a widely used and concise tool used to assess the functional impairment of patients with ME/CFS [51], with 100% indicating normal health and 0% bedriddenness. The Chalder Fatigue Scale (CFQ) evaluates 14 items to measure the impact and severity of fatigue’s physical and mental aspects [52]. The Composite Autonomic Symptom Score-31 (COMPASS-31) is a concise instrument to assess autonomic nervous system dysfunction. It comprises 31 validated items in six domains: orthostatic intolerance, vasomotor, secretomotor, gastrointestinal, bladder, and pupillomotor function, and a total score ranging from 0 to 100 [53].

### Collection and Storage of Supplementary Bio-Samples

If patients provided a sta0ß0000ndard broad consent together with the MECFS-R consent, biospecimens were collected, processed, and stored according to the local central biobank’s standard operating procedures (SOP), and their processing time, type, and number documented within the MECFS-R.

### Statistical Analyses

Statistical analysis was performed using IBM SPSS Statistics 29 (IBM, Armonk, New York, USA) and R version 4.2.1 “Funny Looking Kid” (The R Foundation for Statistical Computing, Vienna, Austria). We employed descriptive statistics and frequency analyses to examine sample characteristics, such as demographics and access to medical care. Fisher’s exact test or Pearson’s χ2 test was employed for comparing categorical variables, while the Wilcoxon rank-sum test was utilized for comparing numeric variables between groups. The significance level was set to α = 0.05.

## RESULTS

### Baseline Characteristics

Here we describe a pilot cohort of 174 patients with ME/CFS enrolled in the MECFS-R from 04/2021 to 03/2023. The cohort had a mean age of 32.6 years (SD 14.9; range 11 – 61). 43/174 (24.7%) patients were children and adolescents, and 136/174 (78.2%) were female. 62/174 (35.6%) patients were recruited at the MCFC, with a mean age of 18.9 years (SD 3.4; range 13 – 28), including 45/62 (72.6%) females. The CFC enrolled 112/174 (64.5%) patients with a mean age of 41.8 years (SD 11.1; range 18 – 62), including 91/112 (81.3%) females (**Figure 1A** and **1B**). The percentage of females was higher among adult patients compared to children and adolescents (81.7% vs. 67.4%, P = 0.050).

**Figure 1.**
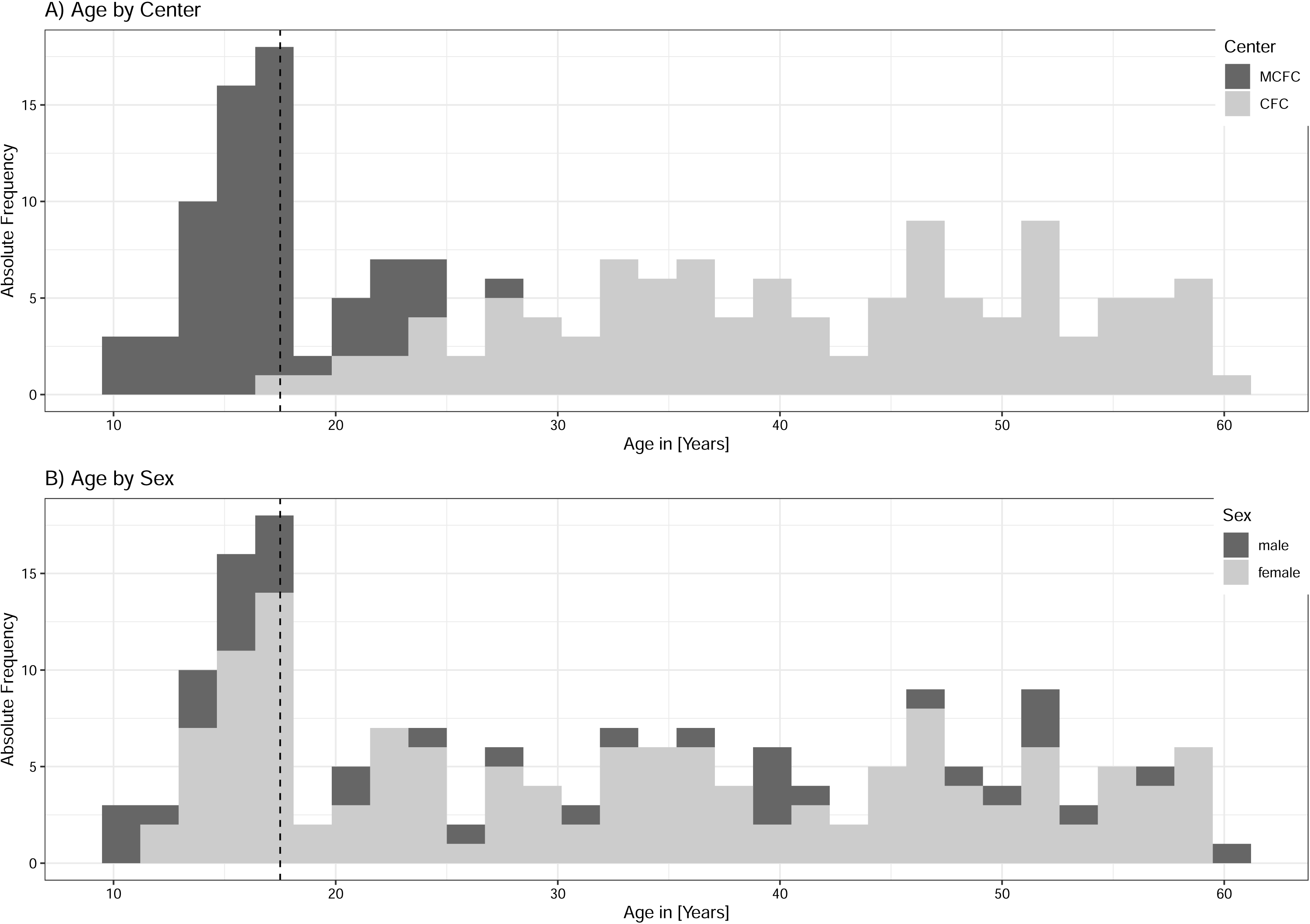
Age Distribution. Histograms show the age distribution of patients included in the MECFS-R depending on the recruiting center (A) and gender (B).

### Participation

At the time of enrollment, 59/158 (37.3%) patients were in school or vocational education, 87/174 (55.1%) were employed, 3/174 (1.9%) were in early retirement, and 9/174 (5.7%) reported no current activity. Following the onset of ME/CFS, **14/152** (9.2%) patients (0/36 children and adolescents vs. 14/116 (12%) adults, P<0.001) were able to maintain their pre-illness participation. **13/152** (8.6%) patients (8/36 (22%) children and adolescents vs. 5/116 (4.3%) adults, P<0.001) participated partially with more than 50% of the pre-illness activity level. **22/152** (14%) (12/36 (33%) children and adolescents vs. 10/116 (8.6%) adults, P<0.001) participated partially with less than 50% compared with the pre-illness level. The majority of patients (**103/152** (68%), including 16/36 (44%) children and adolescents vs. 87/116 (75%) adults, P<0.001) were unable to participate at all in previous education or work.

### Onset of ME/CFS

**160/174** (92.0%) patients (36/43 (83.7%) children and adolescents vs. 125/131 (95.4%) adults, P=0.011) reported an acute viral infection before the onset of ME/CFS. The most frequent confirmed triggers were SARS-CoV-2 in 82/174 (47.1%) patients (78/131 (59.5%) adults vs. 5/43 (11.6%) children and adolescents, P <0.001) and EBV in 19/174 (10.9%) patients (11/43 (26%) children and adolescents vs. 10/131 (7.6%) adults, P=0.012). An influenza virus infection was documented in 2/174 (1.1%) patients (2/43 (4.7%) children and adolescents vs. 0/131 (0.0%) adults, P=0.061). In 5/174 (2.9%) patients (2/43 (4.7%) children and adolescents vs. 3/131 (2.3%) adults, P=0.421) multiple infectious triggers were recalled at the time of disease onset (**Figures 2A** and **2B**). Other confirmed or probable infectious triggers were coxsackieviruses, mycoplasma, Borrelia burgdorferi, respiratory syncytial virus, and Group A streptococcae.

**Figure 2.**
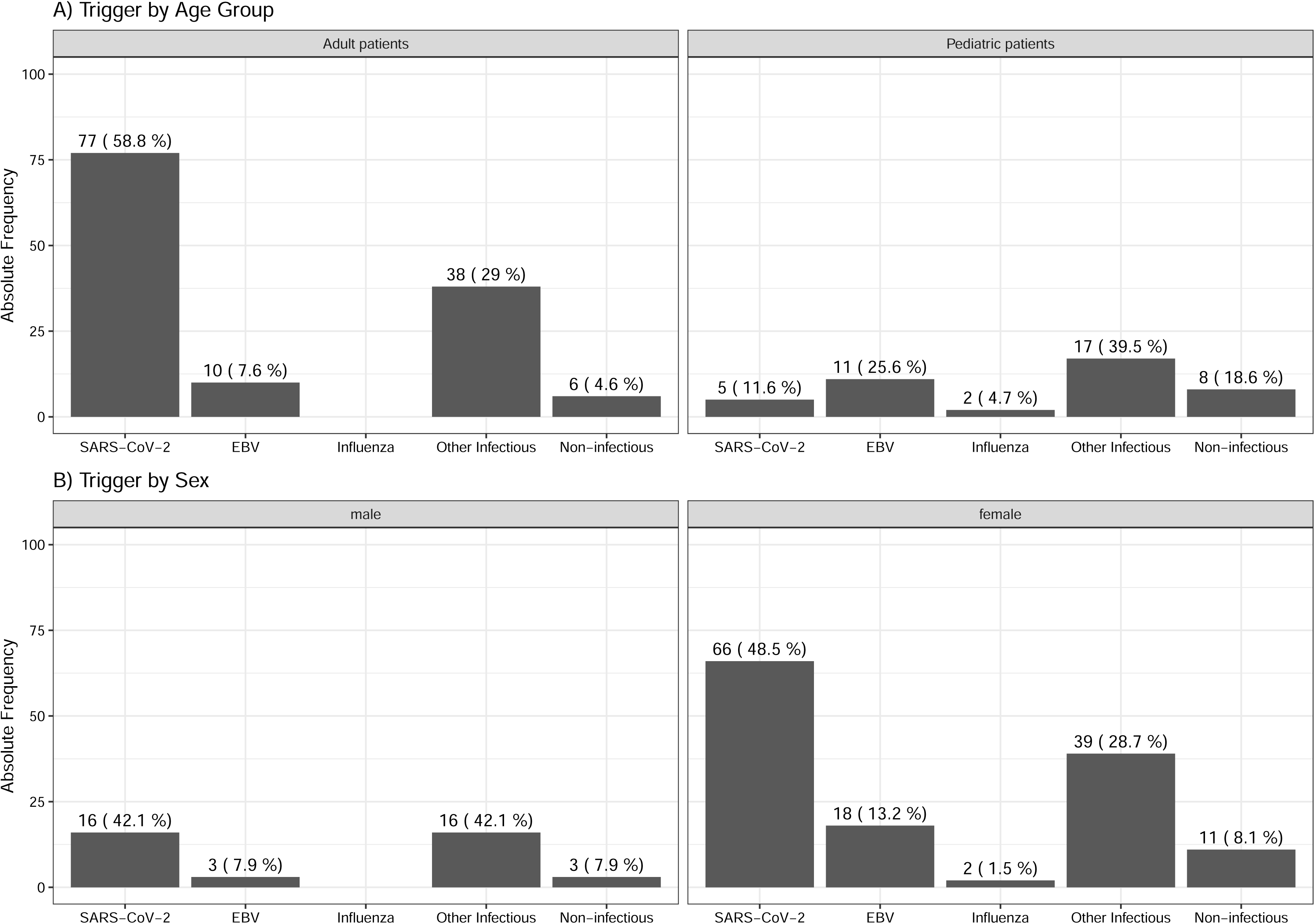
Distribution of ME/CFS Triggers. Bar charts display the absolute frequency and relative percentage of reported ME/CFS triggers by age group (A) and gender (B).

### Diagnostic Criteria and Post-Exertional Malaise

All patients met at least one of the four ME/CFS case definitions (CCC, IOM, CDW-R, PCD-J). Among adult patients tested with the indicated questionnaire 127/129 (98.4%) fulfilled the CCC, 108/108 (100%) the IOM, and 106/108 (98.1%) both. Among children and adolescents tested with the indicated questionnaire, 35/42 (83.3%) fulfilled the CCC, 16/16 (100%) the IOM, 39/39 (100%) the CDW-R, and 16/19 (84.2%) the PCD-J criteria. Most adults fulfilled the CCC (98.1%) and IOM criteria (100%), because until March 2023 only patients fulfilling CCC were included at the CFC. Remarkably, using the DSQ-PEM as a PROM prior to medical assessment at the CFC or MCFC only 139/153 (90.8%) patients scored positive for PEM (21/25 (84.0%) children and adolescents vs. 118/128 (92.2%) adults, P=0.348) while all patients clearly indicated PEM when interviewed by an ME/CFS-experienced physician. Using the DSQ-PEM as a PROM, PEM duration was reported to be 2-3 h by 1/138 (0.7%), 4-10 h by 2/138 (1.4%), 14-24 h by 23/138 (16.6%) (18/117 (15.3%) adults vs. 5/21 (23.8%) children and adolescents), and >24 h by 112/138 (81.1%) (97/117 (82.9%) adults vs. 15/2 (71.4%) children and adolescents) of patients, indicating the majority of patients had long lasting PEM.

### Patient-Reported Outcome Measures

The SF-36 was used to assess HRQoL and showed significantly reduced scores in this ME/CFS cohort across all domains compared to a published healthy German population−based sample (**Figure 3**). Overall, the lowest SF-36 scores were reported for the domains *vitality* and *role physical,* while the highest scores were found for the *mental health* and *emotional role* subscales. Compared to adults children and adolescents displayed significantly higher scores on the domains of mental health (67.9 (SD 16.5) vs. 56.3 (SD 18.8), P = 0.009) and role physical (3.8 (SD 10.0) vs. 0.0 (SD 6.2), P = 0.004) compared to adults. Furthermore, the self-reported health change in the last year was significantly better in children and adolescents (35.6 (SD 32.5) vs. 22.8 (SD 35.6), P = 0.049). The median Bell Score of the cohort was 30.0 (IQR 30.0 – 40.0) (30.0 (IQR 27.5 – 40.0) in children and adolescents vs. 30.0 (IQR 30.0 – 40.0) in adults, P = 0.467), indicating a severely impaired functional status (**Figures 4A** and **4B**). The overall score of the CFQ was 27.6 (SD 3.7). Children and adolescents reported significantly less fatigue than adult patients (24.4 (SD 5.0) vs. 28.0 (SD 3.3), P = 0.022) (**Figures 4C** and **4D**). Most patients (128/174 (73.6%)) who completed the COMPASS-31 suffered from autonomic dysfunction, with moderate symptoms, i.e. a total score between 20 to 40, in 53/128 (41.4%) adults and 61/128 (47.7%) children and adolescents, respectively. The total weighted score of the COMPASS-31 ranged from 2 to 89.9, with a mean of 40.1 (SD 15.9) (**Figure 4E** and **4F**). The COMPASS-31 total scores and sub-scores of orthostatic, gastrointestinal, vasomotor, pupillomotor, secretory, and bladder symptoms are presented in **Table 2**. Children and adolescents had significantly lower scores on the gastrointestinal, bladder, pupillomotor subdomains, and total scores but significantly higher scores for orthostatic intolerance.

**Figure 3.**
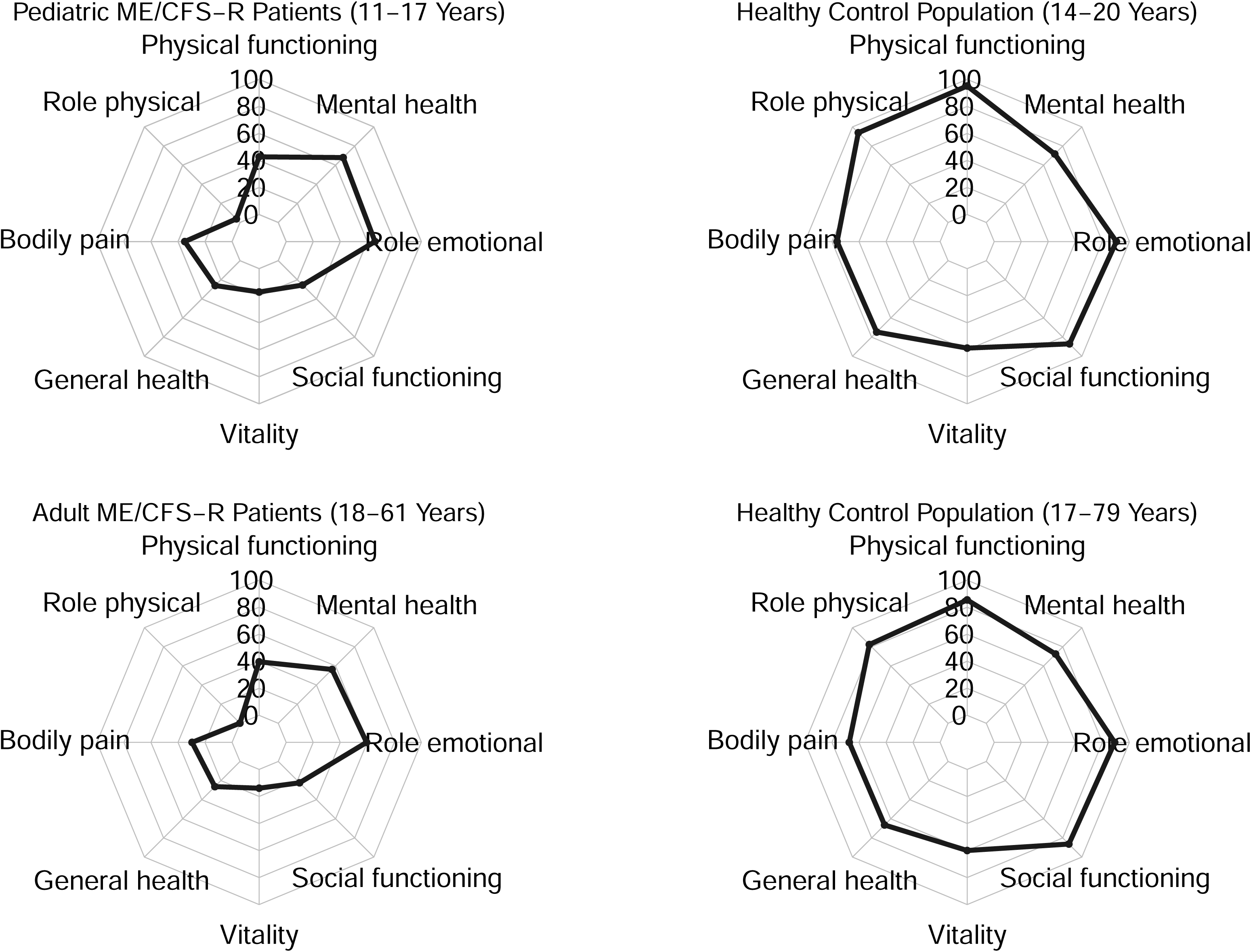
Results from the SF-36 Questionnaire. Spider diagrams display the results form subdomains of the SF-36 questionnaire for pediatric ME/CFS-R patients (age 11-17 years) (top left), adult ME/CFS-R patients (age 18 - 61 years) (bottom left), as well as for largely age-matched historic, healthy control populations aged 14 to 20 years (top right) and 17-79 years (bottom right).

**Figure 4.**
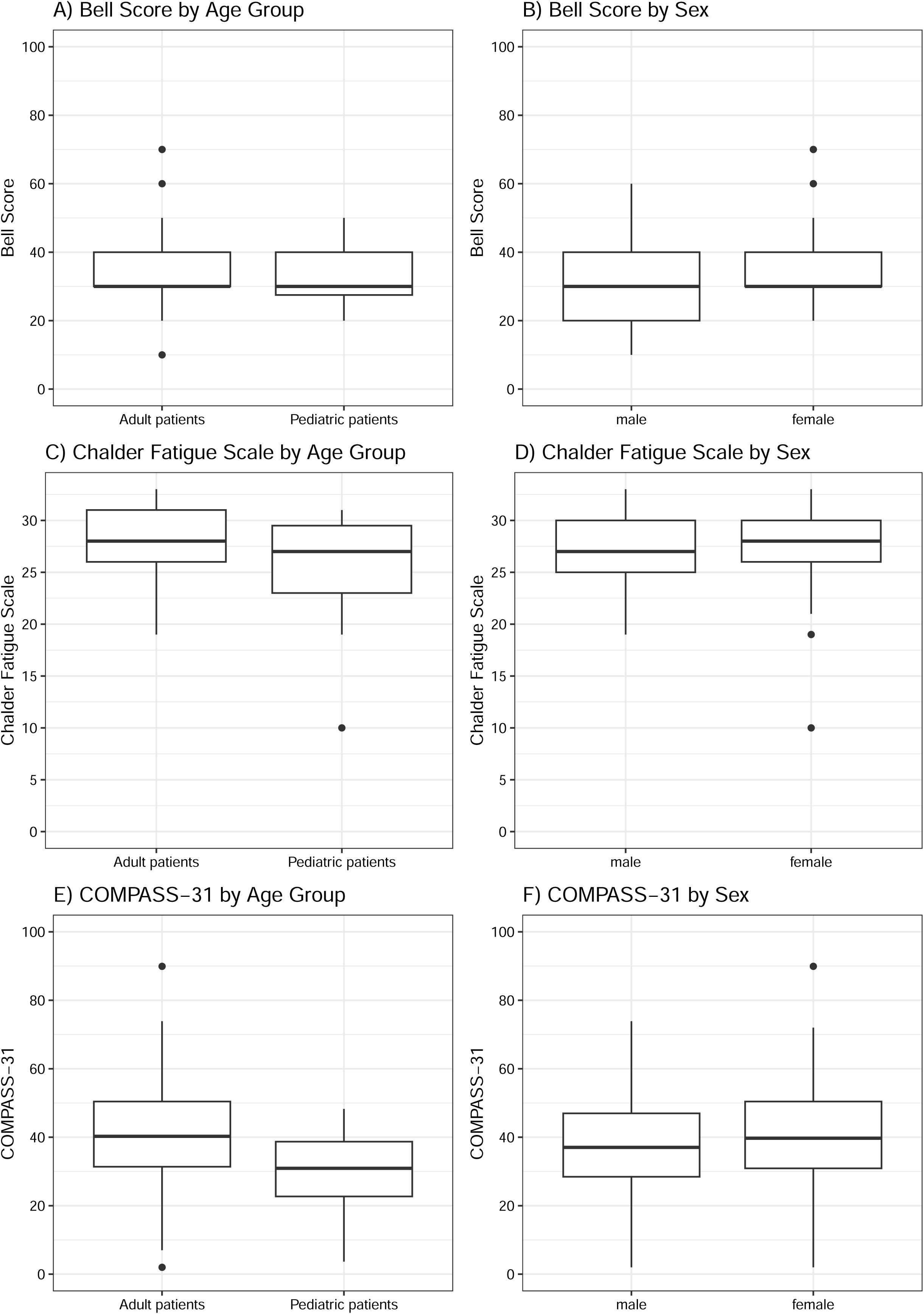
Results from Patient-Reported Outcome Measures. Boxplots display the results from the Bell Score, Chalder Fatigue Scale, and COMPASS-31 questionnaire for children and adolescents *versus* adult patients (A, C, E) and male *versus* female patients (B, D, F).

**Table 2.** Composite Autonomic Symptom Score 31 (COMPASS 31)

| Domain | All<br>(n=128) | Adolescents<br>(n=15) | Adults<br>(n=113) | p-value | Healthy Population<sup>1</sup><br>(n=20) |
| --- | --- | --- | --- | --- | --- |
|  | mean (SD) | mean (SD) | mean (SD) |  | mean (SD) |
| **Orthostatic intolerance** | 13.8 (11.2) | 21.6 (10.0) | 12.8 (11.0) | 0.004 | 5.1 (7.5) |
| **Vasomotor** | 1.0 (1.6) | 0.9 (1.4) | 1.0 (1.6) | 0.986 | 0.3 (0.7) |
| **Secretomotor** | 3.3 (3.0) | 2.0 (2.4) | 3.5 (3.1) | 0.093 | 1.4 (2.1) |
| **Gastrointestinal** | 8.1 (5.1) | 3.5 (3.0) | 8.8 (5.0) | <0.001 | 3.8 (2.9) |
| **Bladder** | 1.2 (1.7) | 0.1 (0.6) | 1.4 (1.8) | 0.002 | 0.3 (0.9) |
| **Pupillomotor** | 2.7 (1.3) | 1.9 (1.0) | 2.9 (1.3) | 0.002 | 0.9 (0.9) |
| **Total** | 40.1 (15.9) | 29.9 (11.7) | 41.5 (15.9) | 0.007 | 11.2 (9.1) |

<sup>1</sup>[53]
Autonomic symptoms were assessed by COMPASS 31 questionnaire, considering the total score (0-100) and the scores of the six subdomains: orthostatic intolerance (0-40), vasomotor (0-5), secretomotor (0-15), gastrointestinal (0-25), bladder (0-10) and pupillomotor (0-5).

## DISCUSSION

### Aim and Structure of the Registry Study

Here, we report on the aims, structure, and implementation of the German ME/CFS registry, including data from a pilot cohort of 174 adult and pediatric patients recruited at the Munich Chronic Fatigue Center for Young People (MCFC) and the Charité Fatigue Center (CFC) in Berlin.

The recent COVID-19 pandemic has resulted in a significant rise in the number of people worldwide experiencing persistent post-viral syndromes, including ME/CFS. Accordingly, scientific and clinical interest and needs in this field are increasing [54]. It is estimated that 19–58% patients with PASC, also known as post COVID-19 condition (ICD-10 U09.9!), meets the diagnostic criteria for ME/CFS [15, 18], representing up to about 50% of patients at healthcare institutions specialized in severe forms of PASC [55]. We recently described ME/CFS following COVID-19 in children as young as 11-14 years, with severe impact on their daily function [56]. Our user-friendly MECFS-R, with its standard dataset, novel questionnaires such as the MBSQ [19], and accompanying information can help PASC teams in developing local standard approaches diagnosing and phenotyping ME/CFS following COVID-19.

Despite the considerable impact on health, participation, and HRQoL of people with ME/CFS as well as significant socioeconomic costs due to this disabling disorder, limited knowledge is available regarding the etiology, risk factors, diagnostic markers, treatment approaches, prognosis, and prevention [57-59]. Research on ME/CFS has been hindered using unsuitable case definitions, relatively small study cohorts, the lack of reliable diagnostic and prognostic biomarkers, and limited funding for research and care [60, 61]. However, generating comprehensive and large-scale routine clinical data in registries can help gain deeper insight into clinical features, pathophysiology, and care options.

To address these issues and facilitate future research on ME/CFS, we developed and implemented the German ME/CFS registry and biobank at two German tertiary care centers specialized in diagnosing and treating ME/CFS in adults, adolescents, and children. This registry aims to harmonize the diagnostic approach to ME/CFS and generate a large, well-characterized study cohort via standardized deep clinical and biological phenotyping.

Instruction manuals and individual training will be provided to future participating centers to support a valid comprehensive standard dataset. We expect to generate knowledge about potential ME/CFS subgroups, natural disease trajectories, and current medical care across all age groups and will provide baseline data for other clinical and translational research.

Previous ME/CFS case definitions often have not required PEM as the cardinal symptom of ME/CFS, resulting in patient cohorts that included non-ME/CFS cases, possibly explaining conflicting research findings [62]. This registry only includes ME/CFS cases defined by diagnostic criteria requiring PEM, including the internationally recommended IOM criteria, the more stringent CCC, and two pediatric CDW-R and PCD-J criteria sets. To ensure a standardized quantitative evaluation of these criteria, the MBSQ was developed as a novel questionnaire with diagnostic algorithms for adults and pediatric patients [45] and is being suggested for use at all participating MECFS-R centers.

In addition, several published PROMs have been selected as important diagnostic tools based on expert recommendations and according to common data elements suggested by the National Institute of Neurological Disorders and Stroke (NINDS) [46]. They address clinical and psychosocial features of ME/CFS such as distinct symptoms, daily function, and HRQoL [63]. We are currently programming all PROMs and additional questionnaires in the REDCap format with mapping to the MECFS-R DIS format to facilitate data capture directly from patients with state-of-the-art data protection measures. The inclusion of a standard dataset as well as additional parameters allows flexible data entry protocols according to the local clinical standards of participating centers.

### Clinical Characterization of Pilot Study Participants

The pilot cohort of 174 patients in this registry included 131 adults as well as 43 adolescents and children with ME/CFS. The female predominance and age peaks observed in this cohort are well-known for ME/CFS [64, 65]. The youngest patient in our cohort was 11 years old, in line with a lower prevalence of ME/CFS in childhood compared to adolescence and adulthood [66].

Almost all adult patients fulfilled both the CCC and IOM criteria. The proportion of study participants who met the evaluated case definition was 100% for IOM and CDW-R and approximately 89% and 83% for the more stringent PCD-J and CCC, respectively. According to the medical interview, not all patients with physician-validated PEM fulfilled the PEM criteria when using the DSQ-PEM as a PROM. This is congruent with our clinical experience demonstrating that self-assessment of PEM and its duration can be difficult, especially in young patients and patients who largely avoid PEM by consequent pacing. The newly established, age-adapted MBSQ, together with the DSQ-PEM, thus help in assessing PEM and diagnosing ME/CFS [45] but cannot replace a detailed personnel medical interview.

ME/CFS is known to be most commonly triggered by an acute viral disease, with a significant impact of the COVID-19 pandemic on ME/CFS prevalence [67]. Accordingly, a predominance of SARS-CoV-2 was identified in adults (59.5%) and EBV in pediatric patients (26%) in our first cohort. Non-infectious triggers are most likely underrepresented since both recruiting centers are focusing on post-infectious ME/CFS as immunological departments [55, 56, 68]. However, the registry allows a very precise documentation of triggering events including clinical and laboratory data from the time of initial symptoms, and therefore facilitates a stratification of study participants along confirmed *versus* probable and self-reported triggers.

Notably, only a minority of study participants were able to work, and more than half of the children and adolescents were not able to participate in school. This was in line with previous studies reporting a worrying impact of ME/CFS on education and social participation [69]. The physical and social functioning of MECFS-R participants was severely reduced as indicated by low Bell and SF-36 scores, while higher scores were found for emotional role functioning and psychological well-being [70, 71]. This aligns with earlier reports indicating that the HRQoL of patients with ME/CFS compared to other chronic diseases is severely compromised, mainly due to physical impairment [55, 56, 68, 72] Moreover, in support of published results [73], MECFS-R participants suffered from significant autonomic dysfunction as indicated by high COMPASS-31 scores. We recommend the Bell score, SF-36, and COMPASS-31 as standard measures for clinical phenotyping to facilitate both local medical care as well as future studies with secondary use of MECFS-R data.

### Strengths and Limitations

To our knowledge, this is the first multicenter registry collecting cross-age routine clinical data and information on biosamples from ME/CFS patients diagnosed by trained staff and in a standardized manner at specialized tertiary care centers, with obligatory quantification of ME/CFS symptoms and detailed assessment of PEM as an essential diagnostic criterion.

To date, few registries for ME/CFS exist with different scopes and selection criteria. The UK biobank includes patients diagnosed with ME/CFS by primary care physicians and complies with the CCC and/or the CDC-1994 (“Fukuda”) criteria [74], and the YOU+ME registry relies on self-report. Both approaches support collecting large-scale data but might face the risk of false diagnoses and lack much of the detailed clinical information provided by the MECFS-R. The MECFS-R offers a comprehensive dataset with more than 10,000 variables per patient for secondary use in future clinical and translational studies, including standardized data on clinical phenotypes, patient journeys, and impact on daily life.

A strength of the MECFS-R is the collection of routine data which means that neither the patient nor the treating physician must make an extra effort to participate, except for the informed consenting procedure. Furthermore, different levels of data complexity can be chosen by the participating centers, and datasets can be adapted to local clinical care protocols.

We provide a selected core dataset from a pilot group of study participants as an example which well aligns with published data from other cohorts. Especially in pediatrics, the MECFS-R is expected to contribute significant novel evidence in many aspects of this complex disease.

However, since routine data are collected, follow-up visits documented in the MECFS-R do not follow strict protocols as in prospective cohort studies. As a second limitation, the quality and quantity of individual data sets might differ depending on the level of training and resources available for documentation at the participating hospitals or private practices. However, subgroup analyses will allow for interpretation without bias and even small data sets from many patients might contribute important information. Finally, the pilot group of patients presented here is relatively small and not representative, but provided important data to validate the comprehensive MECFS-R concept.

## Conclusion

We here first report on a multicenter German ME/CFS registry study, which collects comprehensive, standardized data on clinical features and biospecimens from adults, adolescents, and children. The MECFS-R team consented to a large set of core diagnostic measures and offers specific training to members of future participating centers. The inclusion of patients with well-defined ME/CFS and obligatory PEM, together with detailed information on clinical and laboratory findings as well as collected biosamples, is expected to significantly enhance clinical and translational research on ME/CFS and thereby improve medical care for affected patients of any age in Germany and beyond.

## Abbreviations

CCC: Canadian Consensus Criteria
CDW-R: Clinical Diagnostic Worksheet by P.C. Rowe and colleagues
CFC: Charitè Fatigue Center
CFS: Chronic Fatigue Syndrome
COMPASS-31: Composite Autonomic Symptom Score-31
COVID-19: Coronavirus Disease 2019
CRF: Clinical report form
DIFUTURE: Data Integration for Future
DIS: Data Integration System
EBV: Epstein-Barr virus
HRQoL: Health-related quality of life
ICD-10: International Classification of Diseases
IM: Infectious Mononucleosis
IOM: Institute of Medicine
MCFC: Munich Chronic Fatigue Center for Young People
MBSQ: Munich Berlin Symptom Questionnaire
ME: Myalgic Encephalomyelitis
MECFS-R: ME/CFS-Registry
NINDS: National Institute of Neurological Disorders and Stroke
PASC: Post-acute sequelae of SARS-CoV-2 infection
PBMC: Peripheral Blood Mononuclear Cells
PEM: Post-Exertional Malaise
PCC: Post COVID-19 condition
PCD-J: Pediatric Case Definition by L.A. Jason and colleagues
PROM: Patient-reported outcome measure
SARS-CoV-2: Severe acute respiratory syndrome coronavirus type 2
SD: Standard deviation
SF-36: Short form-36 health survey

## DECLARATIONS

### Author Contributions

R.P., H.H., M.H., K.W., K.G., C.S., and U.B. designed the study. H.H., R.P., M.H., K.W., Q.D., C.C., C.W., S.S., K.W., H.F., A.L., A.K., Y.M., H.Z. and C.H. acquired data and investigated patients. H.H., R.P., and L.M. analyzed the data. H.H., R.P., L.M., C.C., D.H., and U.B. drafted and reviewed the manuscript. K.G., C.S. and U.B. supervised this work. All authors discussed the results, commented on the draft, and consented to the final manuscript.

### Institutional Review Board Statement

The study was approved by the local ethics committee at TUM (116/21 S) and Charitè (EA/006/22) and conducted in accordance with the declaration of Helsinki.

### Informed Consent Statement

Patients and legal guardians of patients younger than 18 years gave written and informed consent prior to inclusion into the MECFS-R.

### Data Availability Statement

The data presented in this study are available from the corresponding author, upon reasonable request.

